# Impact of a geriatric emergency management nurse on thirty-day emergency department revisits: a propensity score matched case-control study

**DOI:** 10.1101/2024.12.30.24319195

**Authors:** Nathalie Germain, Rawane Samb, Émilie Côté, Annie Toulouse-Fournier, Joanie Robitaille, Stéphane Turcotte, Michèle Morin, Martyne Audet, Laetitia Bert, Josée Rivard, Audrey-Anne Brousseau, Lucas B. Chartier, Nadia Sourial, France Légaré, Holly O. Witteman, Clémence Dallaire, Chantal Kroon, Patrick Archambault, the LEARNING WISDOM investigators, Network of Canadian Emergency Researchers

**Affiliations:** Centre de recherche intégrée pour un système apprenant en santé et services sociaux, Centre intégré de santé et services sociaux de Chaudière-Appalaches, Lévis, Québec, Canada; Faculty of Medicine, Université Laval, Québec, Québec, Canada; VITAM - Centre de recherche en santé durable, Québec, Québec, Canada; Département de médecine familiale et médecine d’urgence, Université de Sherbrooke, Sherbrooke, Canada; Department of Emergency Medicine, University Health Network, Toronto, Ontario, Canada; Département de gestion, d’évaluation et de politique de santé, École de Santé Publique, Université de Montréal, Montréal, Canada; Department of Family Medicine and Emergency Medicine, Université Laval, Québec, Québec, Canada; Canadian Association of Emergency Physicians, Ottawa, Ontario, Canada

**Author notes:** Corresponding author: Nathalie Germain.

**Keywords:** Geriatrics, comprehensive geriatric assessment, emergency department revisits, propensity score analysis

## Abstract

**Objectives:** Geriatric Emergency Management (GEM) nurses aim to reduce adverse outcomes by addressing unique needs of older adults seen in emergency departments (EDs), but evidence to demonstrate their impact on ED care transitions is mixed. We evaluated the impact of implementing a GEM nurse model in a local ED on thirty-day revisits using propensity score matching to control for relevant patient characteristics.

**Methods:** A case-control design was used to analyze older adult patients who were triaged to a stretcher at an ED in Lévis, Québec, from October 2018 to September 2019. We used propensity score matching to compare patients who received the GEM nurse intervention with control patients who did not receive the intervention. This intervention involved a targeted geriatric ED assessment including history, physical exam, chart review, communication with caregivers and home care services, and the creation of an intervention and care transition plan to support safe discharge from the ED. We followed both the EQUATOR network’s brief guidelines for reporting a propensity score analysis and the STROBE guideline.

**Results:** Out of 21,024 patients visiting the ED over a one-year period, 7,952 were eligible for analysis, with pre-matching differences showing GEM patients were older and more frequent ED users. Propensity score matching resulted in 724 patients with no significant differences in baseline characteristics between groups. Using a Cox regression analysis, we found a non-significant 6% decrease in the risk of ED revisit within 30 days for the GEM group (HR = 0.94, *p* = .692).

**Conclusions:** The GEM nursing intervention targeting better care transition plans personalized to the needs of each patient did not significantly impact thirty-day revisits to the ED. Further work is needed to determine the most effective specific components of such interventions to maximize future positive impact on the care transitions of older patients.

**Clinician’s capsule:** *What is known about the topic?:* Geriatric emergency management (GEM) nurses heterogeneously contribute to reducing emergency department revisits among older adults but may improve the quality of care.

*What did this study ask?:* How would the implementation of a Geriatric Emergency Management (GEM) nurse intervention in a local emergency department (ED) impact 30-day revisits among older adults?

*What did this study find?:* Despite not reaching statistical significance, we observed a 6% reduction in revisit rates.

*Why does this study matter to clinicians?:* We should refine and support GEM nurse practices, along with shifting outcome measures from service-level metrics to patient-centered metrics like quality of life and symptom burden.

## Introduction

The emergency department (ED) often lacks the specialized multidisciplinary care needed for older adults, who often experience complex comorbidities and frailty, making them the highest users of health-care services [1]. Older adults also have the highest ED use, high rates of ED revisits, and the longest length of ED stays, despite the ED being an unfriendly environment for geriatric patients [2]. Once treated and discharged, they may be more at risk to revisit the ED because they may require ongoing and frequent medical attention.

Hospital-based geriatric interventions generally have little overall effect on ED utilization, whereas outpatient, primary care or home care assessment (including geriatric assessment and management and case management) do reduce ED utilization [3]. Geriatric Emergency Management (GEM) nurses identify, assess, and liaise with older adults to identify appropriate services and practices to reduce adverse outcomes [4]. This approach may meet both the unique health and functional needs of older adults who present to the ED.

In a systematic review, GEM nurses heterogeneously contributed to reducing ED revisits [4], with some studies indicating a reduction, [5] others no effect, [6] and one found an increase in return ED visits [7]. Models of care employed in ED service delivery and enhancement for older adults are trending toward a multidisciplinary approach (e.g., SWAT [8], TREAT [9], ASET [10] and GEDI WISE [11]). These models show heterogeneous benefits, and some are weakened by the absence of convincing evaluative research outcomes. Our objective was to conduct a quasi-experimental design to document the impact of implementing a GEM nurse model in our local ED.

## Method

### Design and Setting

We developed a propensity score matched case-control study of older adult patients who were triaged to a stretcher at the ED at [*hospital name blinded*], in south-eastern Quebec, Canada between 2018-10-01 and 2019-09-30. Early revisits were defined as revisits within 30 days of discharge from the index visit. This project was authorized by the Research ethics board of [*blinded*] with a waiver of informed consent. We followed the brief guidelines for reporting a propensity score analysis and the STROBE guideline [12,13].

### Intervention and comparison

As part of a context-adapted Acute Care for Elders (ACE) intervention developed for the LEARNING WISDOM project [14] and the International ACE Collaborative [15], the GEM nurse intervention was the main intervention implemented during the study.

The intervention provided by a single GEM nurse was a targeted geriatric assessment detailed in Appendix A. Patients were not randomized to the intervention. Target patients were complex cases with physical and social limitations, but whose medical condition seemed stable enough to consider being discharged back to the community after their visit to the ED. Controls were time-concurrent patients who did not receive the GEM nurse intervention. Because patients seen by the GEM nurse had distinct characteristics, our control group was sampled on relevant characteristics with propensity-score matching to isolate for the effect of the intervention.

### Participants

We defined a patient’s index visit as the first visit to the ED made by the patient during the yearlong study period (October 1st, 2018, to September 30th, 2019). Inclusion criteria included: (1) aged ≥ 65 years at the time of the index visit; (2) received care while on a stretcher in the ED observation unit during the index visit; and (3) were discharged home or admitted to the hospital after this first visit. Patients were excluded if they died while in hospital, or if they were transferred to long-term care from hospital.

### Data collection

Data were collected using a clinical administrative database (*Med-GPS*, MediaMed Technologies, Mont-Saint-Hilaire, Quebec). For matching, we collected covariates likely associated with an early ED revisit and that were not influenced by the GEM nurse intervention, to reduce bias. We collected the patient’s age, biological sex, and whether or not they had a family doctor. Then, for the ED visit, we recorded the patient’s provenance (home, care home, referred from a clinic, transferred from hospital), mode of arrival (ambulance or walk-in), major diagnostic category using the International Classification of Diseases, 10th Revision, with Canadian Enhancements [ICD-10-CA], autonomy after triage (stretcher, or waiting room), time of visit (during or not during typical working hours, which were weekdays from 8AM to 6PM), orientation after the visit (admitted to the ward, or discharged home), and whether or not the patient was a frequent user of the ED. We defined frequent users as patients who consulted the ED at least 5 times in the 12 months before the index visit [16]. We also documented diagnoses or presentations associated with geriatric syndromes that would be prioritized by the GEM nurse in her practice (e.g., delirium or confusion, trauma or falls, genitourinary problems).

### Statistical analysis

Data were analyzed with R (Version 4.3.1). Quantitative variables were summarized using their mean and standard deviation if normally distributed, and with the median and interquartile range if otherwise. Qualitative variables were summarized by their frequency distributions. We compared patient characteristics according to whether they were seen or not seen by the GEM nurse. To analyze the effect of the GEM nurse on the overall risk of an early ED revisit, we also report the proportion of patients who returned to the ED within 30 days based on these two groups in the matched sample.

### Propensity score matching

We used the *MatchIt* package in R (version 4.5.5) [17] to perform propensity score matching using logistic regression with the optimal matching method [18] without replacement and matching in a 1:1 ratio. Our chosen model used the average treatment effect as its target estimand [19].

Variable selection was based on clinical importance and available information in the database. Using propensity score analyses necessitate two methodological assumptions: 1) assignment to the experimental arm or the control arm is independent of the potential outcomes conditional on the observed baseline covariates, and 2) every subject had a nonzero probability to receive either the experimental condition or control. We believe that we adequately met this first assumption and that all patients included in propensity score matching could have been selected as candidates to receive the GEM nurse intervention [20]. We used the independent samples *t*-test, Fisher’s exact test, the standardized mean difference, and the risk difference to demonstrate the comparability of baseline characteristics in the GEM nurse group and the control group. There were no missing values for any of the variables included in the propensity score matching model.

### Survival analysis

A Kaplan-Meier model with right-censoring was used to compare the survival curves of patients in the GEM nurse group versus the control group. Each patient had an observation period of thirty days. For each patient, the observation period started at the moment of discharge from the ED to the community. If a patient was admitted to hospital during their index visit, the observation period started at the moment of discharge from hospital to the community. In this model, if patients did not experience a revisit within 30 days of the index visit, they were right censored at day thirty-one. Any ED revisit within 30 days of the index visit was considered an event. We defined survival time as the patient’s number of days within their community since discharge on the initial ED index visit. Any patients that died in community were censored from the analysis on the day of their death. We hypothesized that the GEM nurse intervention would be successful if patients experienced more days in their communities. We then conducted a univariate Cox proportional hazards model to obtain an unbiased estimate of the relative change in the hazard of an early revisit to the emergency department attributable to the GEM nurse intervention.

## Results

### Participant characteristics

Eligible patients retrieved from the database numbered 7,952, representing approximately 35% of the 22,570 patients (See Figure 1). Important pre-matching differences in characteristics were observed for patients seen by the GEM nurse: frequent ED users were more likely to have been seen by the GEM nurse (24% in the intervention group versus 5% in the control group) and patients seen by the GEM nurse were 8 years older on average. Nearly all patients had a family doctor on file, and most were women. Thirteen patients died in their communities within thirty days of ED discharge, one of which was seen by the GEM nurse.

**Figure 1.**
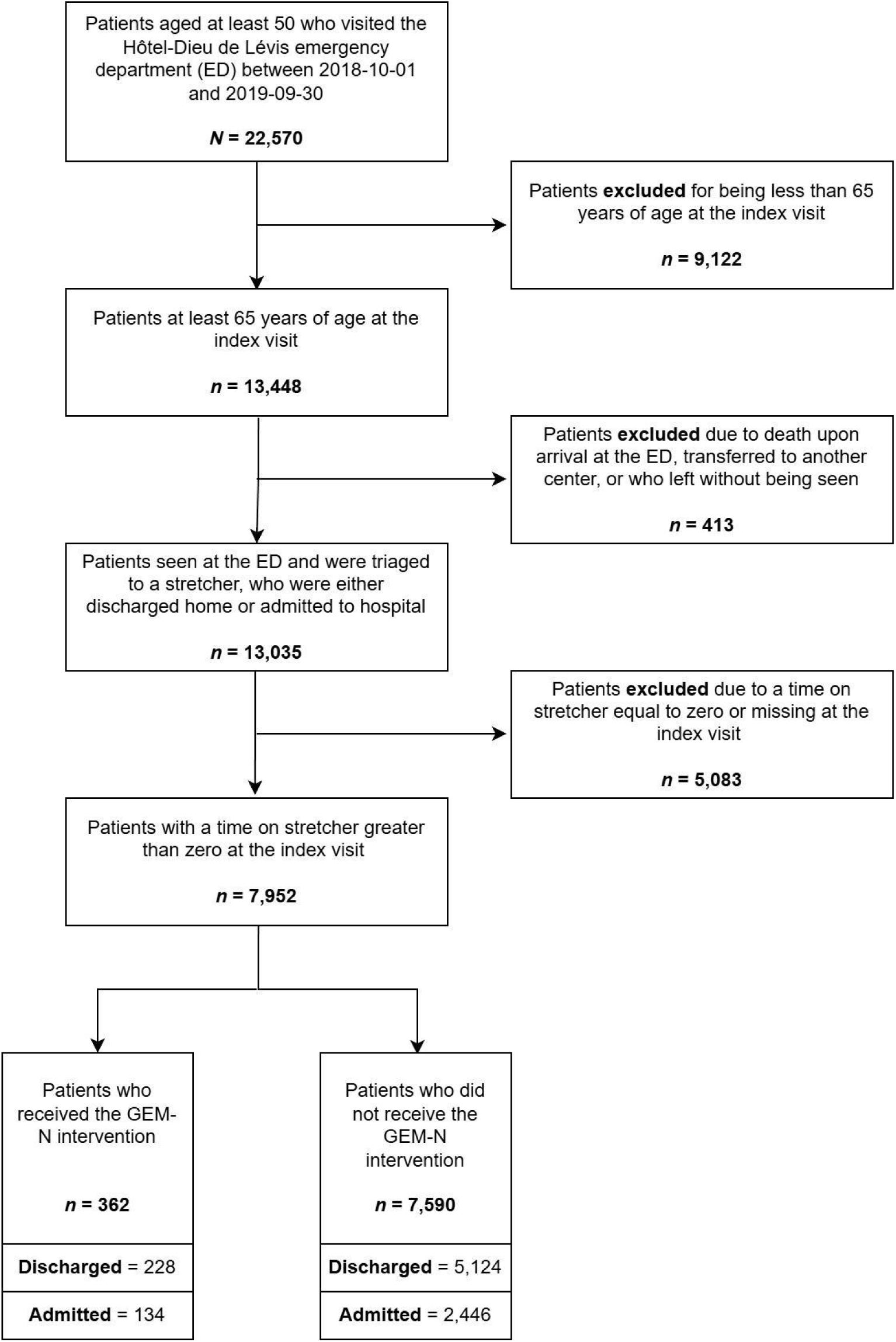
Recruitment flowchart of patient selection and outcomes at the Hôtel-Dieu de Lévis ED between 2018-10-01 and 2019-09-30

### Matching and survival analysis

The propensity score matching yielded two groups each with 362 patients for a total of 724 patients (See Table 1). Matching variables were patient age, patient sex, arrival method, their level of autonomy after triage, triage priority (CTAS) [21], provenance, the date of the index visit (week or weekend), the major diagnostic category, and whether the patient was a frequent user of the emergency department. Major diagnostic category and primary complaint or diagnosis could not both be used as matching variables due to multicollinearity. Adequate balance of baseline covariates was achieved as no differences in means or proportions were statistically significant. In total over thirty days, 179 patients revisited the ED, 87 in the GEM nurse group and 92 in the matched control group.

**Table 1.**
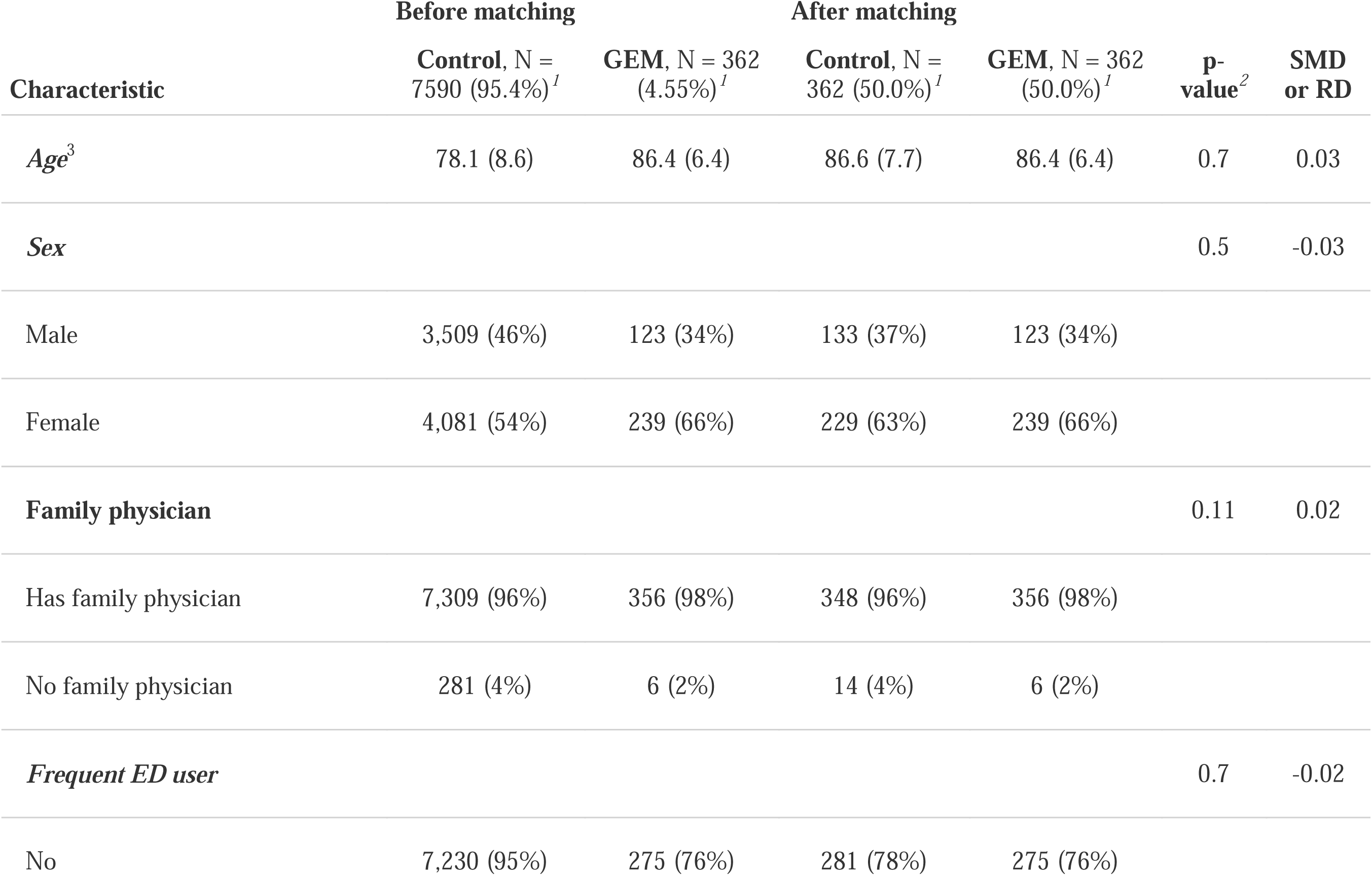
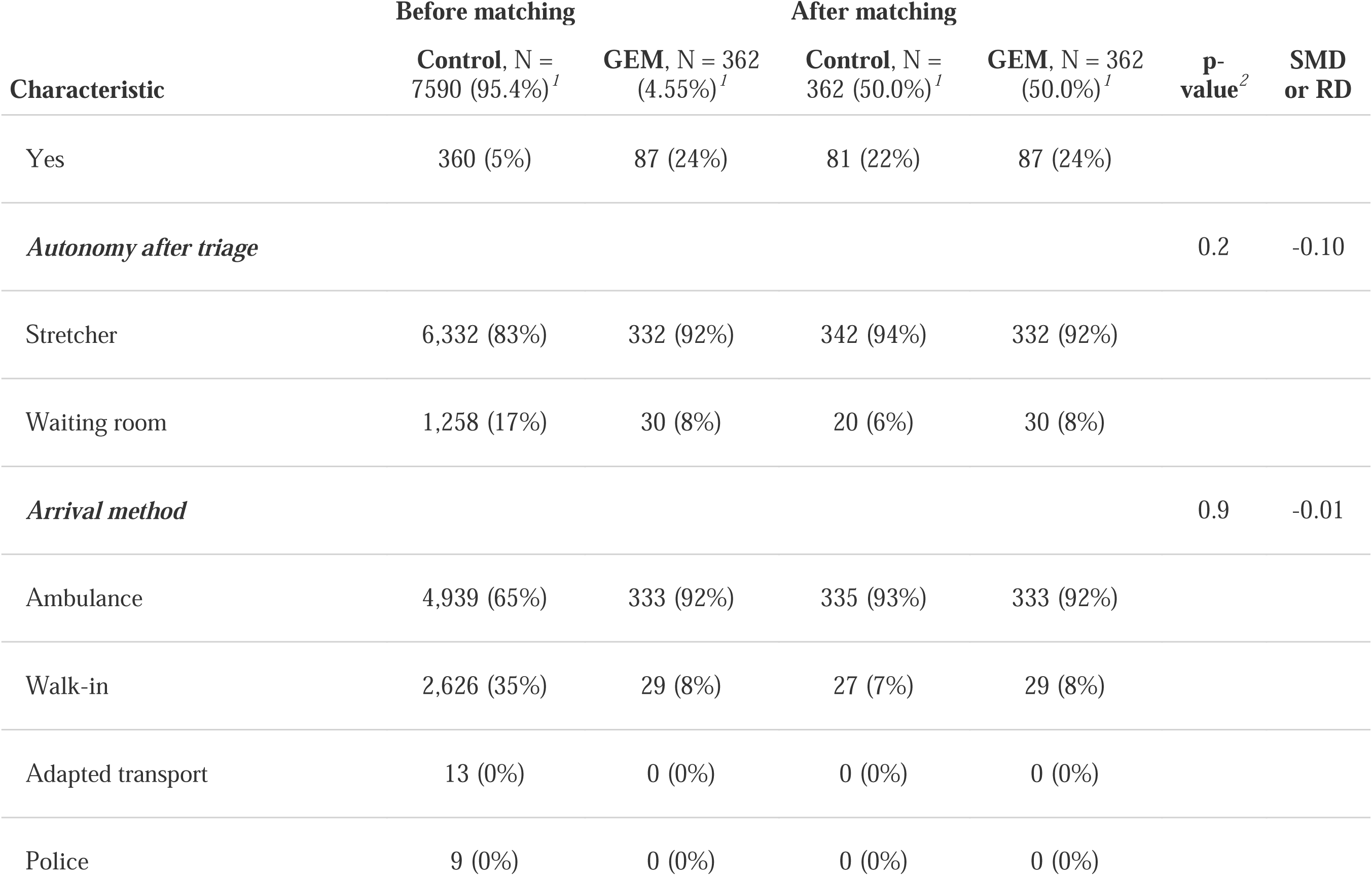
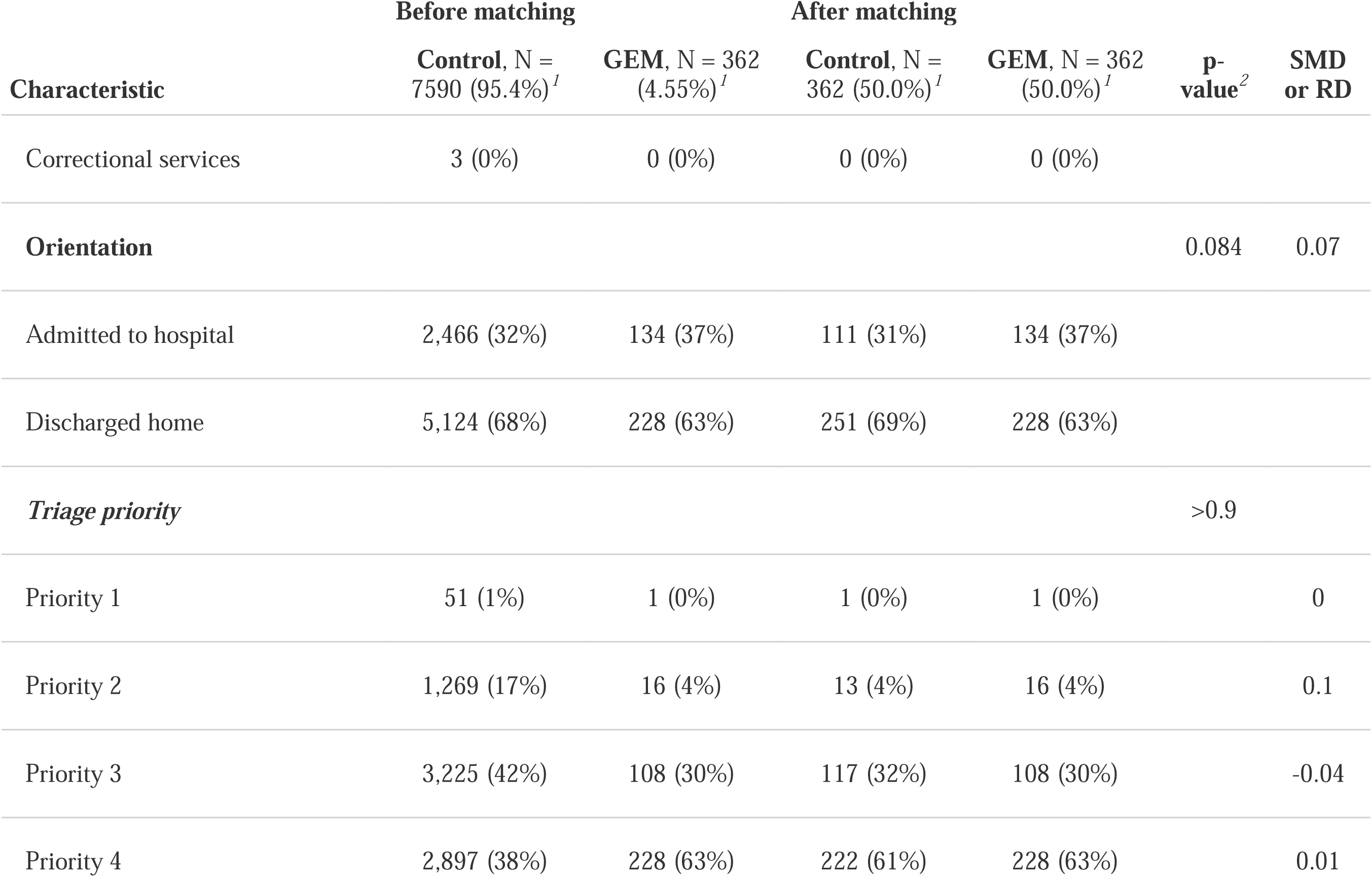
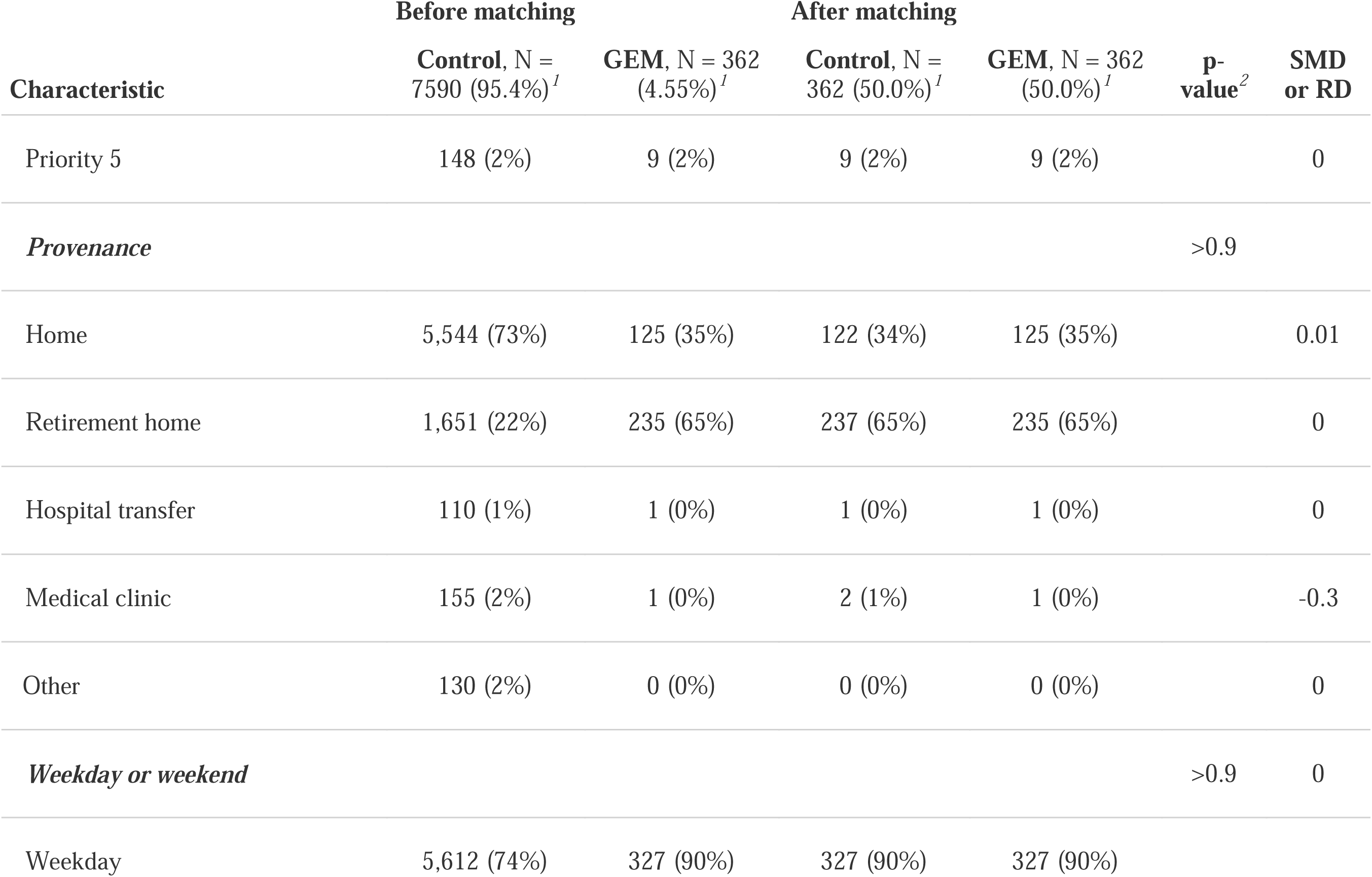
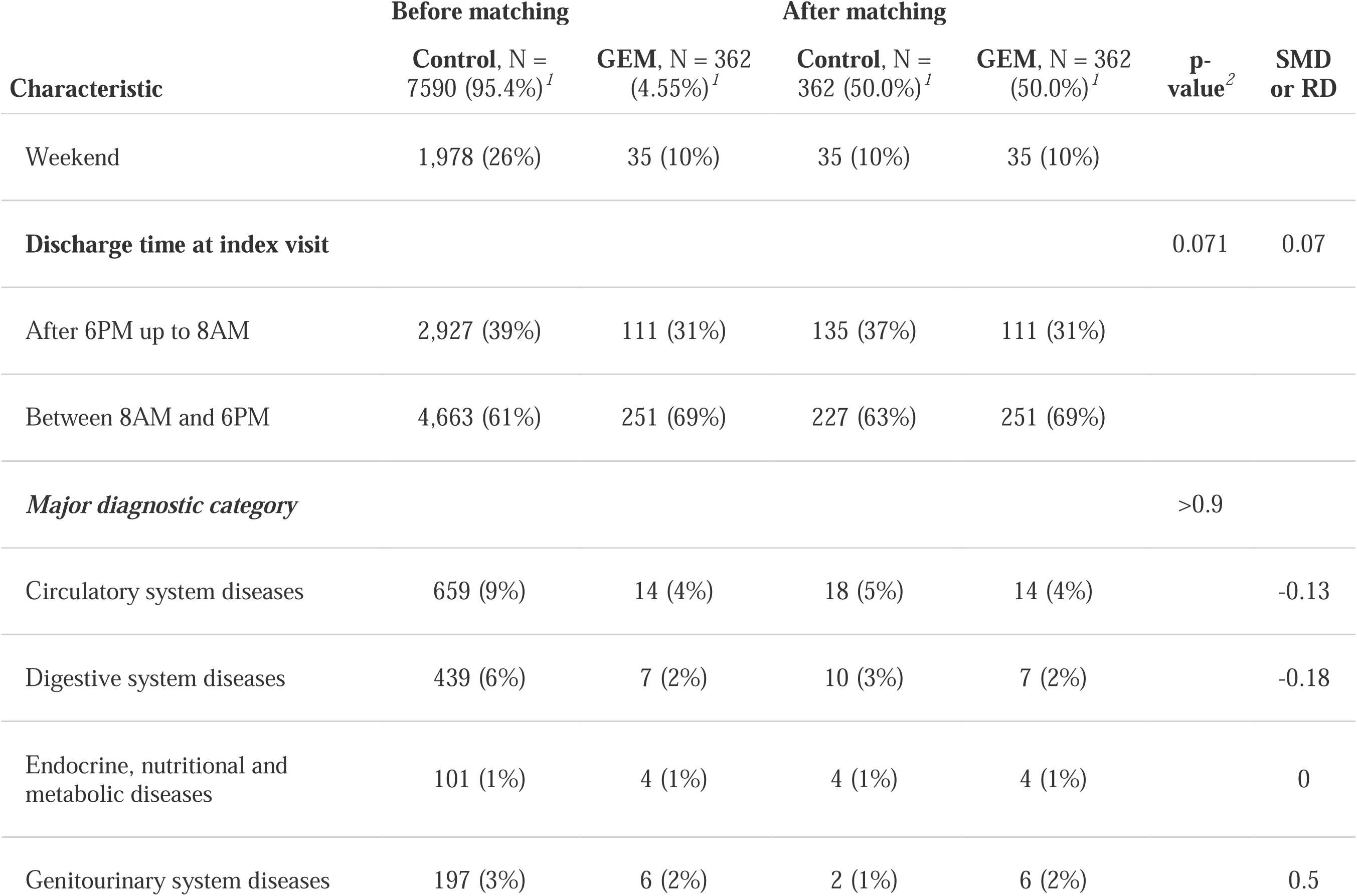
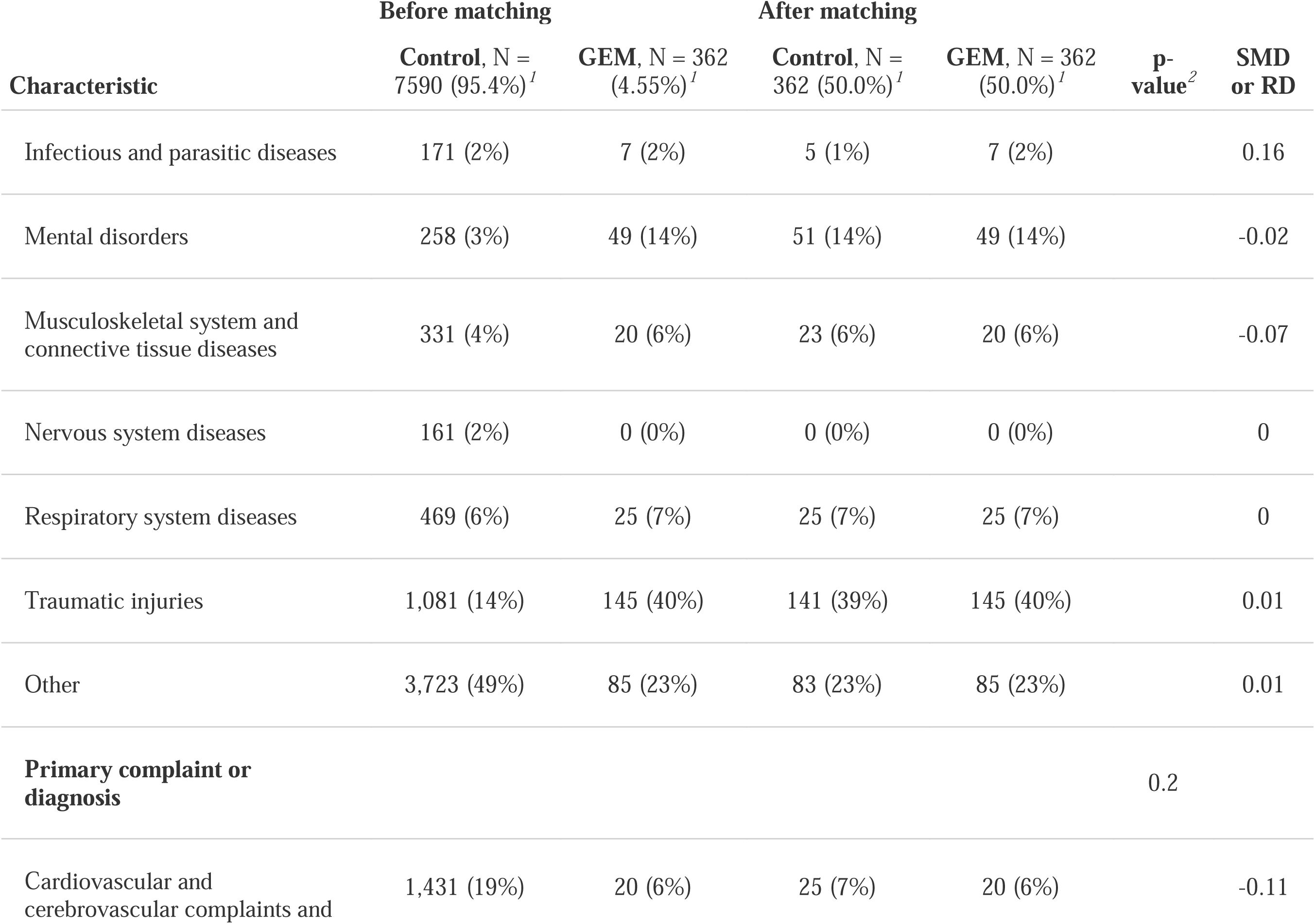
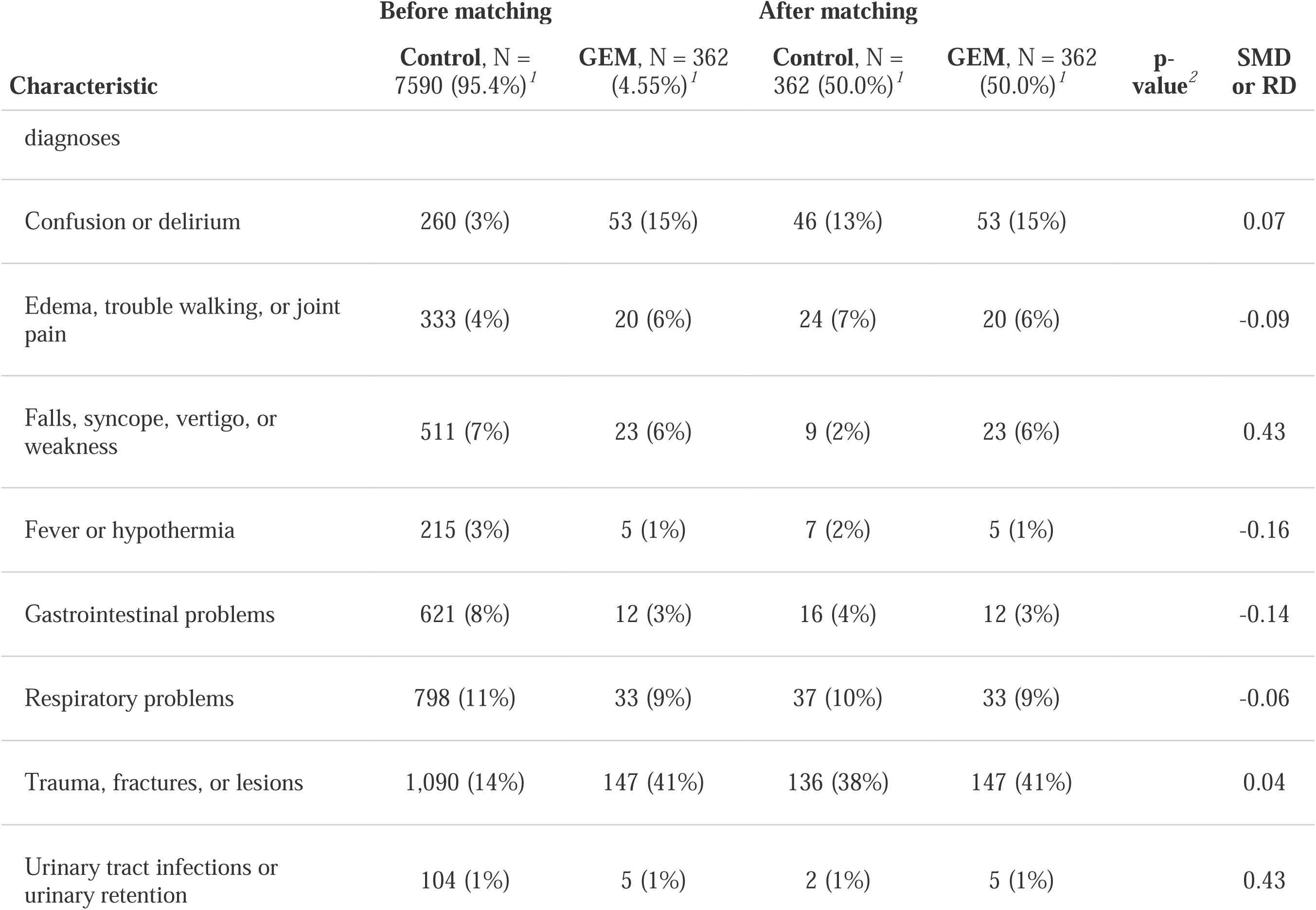
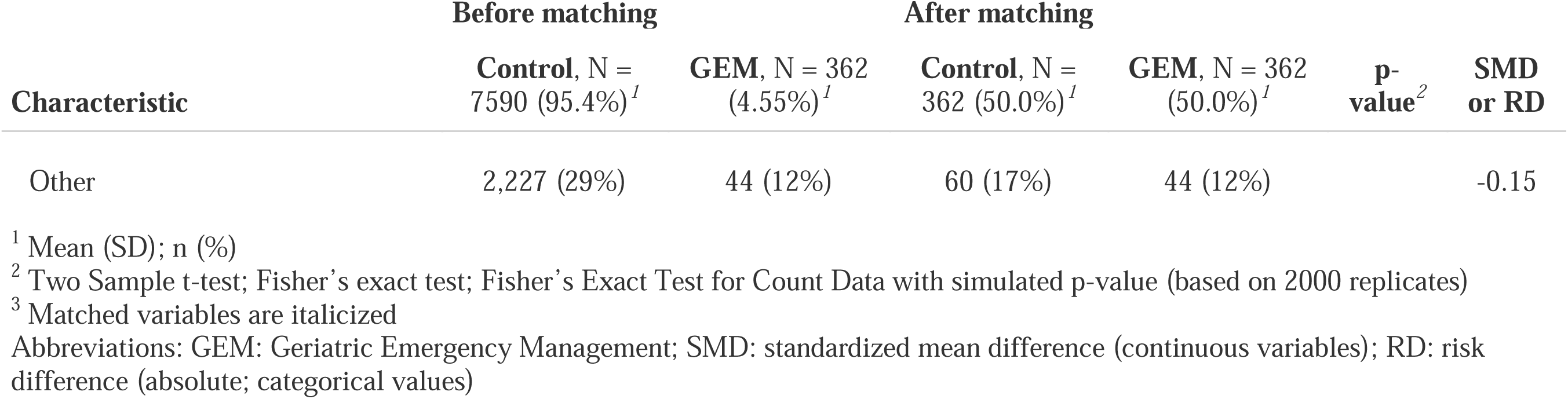
Demographic characteristics of all patients before and after propensity score matching.

| Characteristic | Before matching |  | After matching |  | p-value<sup>2</sup> | SMD or RD |
| --- | --- | --- | --- | --- | --- | --- |
|  | Control, N = 7590 (95.4%)<sup>1</sup> | GEM, N = 362 (4.55%)<sup>1</sup> | Control, N = 362 (50.0%)<sup>1</sup> | GEM, N = 362 (50.0%)<sup>1</sup> |  |  |
| *Age*<sup>3</sup> | 78.1 (8.6) | 86.4 (6.4) | 86.6 (7.7) | 86.4 (6.4) | 0.7 | 0.03 |
| *Sex* |  |  |  |  | 0.5 | -0.03 |
| Male | 3,509 (46%) | 123 (34%) | 133 (37%) | 123 (34%) |  |  |
| Female | 4,081 (54%) | 239 (66%) | 229 (63%) | 239 (66%) |  |  |
| **Family physician** |  |  |  |  | 0.11 | 0.02 |
| Has family physician | 7,309 (96%) | 356 (98%) | 348 (96%) | 356 (98%) |  |  |
| No family physician | 281 (4%) | 6 (2%) | 14 (4%) | 6 (2%) |  |  |
| *Frequent ED user* |  |  |  |  | 0.7 | -0.02 |
| No | 7,230 (95%) | 275 (76%) | 281 (78%) | 275 (76%) |  |  |
|  | Control, N = 7590 (95.4%)<sup>1</sup> | GEM, N = 362 (4.55%)<sup>1</sup> | Control, N = 362 (50.0%)<sup>1</sup> | GEM, N = 362 (50.0%)<sup>1</sup> |  |  |
| Yes | 360 (5%) | 87 (24%) | 81 (22%) | 87 (24%) |  |  |
| *Autonomy after triage* |  |  |  |  | 0.2 | -0.10 |
| Stretcher | 6,332 (83%) | 332 (92%) | 342 (94%) | 332 (92%) |  |  |
| Waiting room | 1,258 (17%) | 30 (8%) | 20 (6%) | 30 (8%) |  |  |
| *Arrival method* |  |  |  |  | 0.9 | -0.01 |
| Ambulance | 4,939 (65%) | 333 (92%) | 335 (93%) | 333 (92%) |  |  |
| Walk-in | 2,626 (35%) | 29 (8%) | 27 (7%) | 29 (8%) |  |  |
| Adapted transport | 13 (0%) | 0 (0%) | 0 (0%) | 0 (0%) |  |  |
| Police | 9 (0%) | 0 (0%) | 0 (0%) | 0 (0%) |  |  |
|  | Control, N = 7590 (95.4%)<sup>1</sup> | GEM, N = 362 (4.55%)<sup>1</sup> | Control, N = 362 (50.0%)<sup>1</sup> | GEM, N = 362 (50.0%)<sup>1</sup> |  |  |
| Correctional services | 3 (0%) | 0 (0%) | 0 (0%) | 0 (0%) |  |  |
| **Orientation** |  |  |  |  | 0.084 | 0.07 |
| Admitted to hospital | 2,466 (32%) | 134 (37%) | 111 (31%) | 134 (37%) |  |  |
| Discharged home | 5,124 (68%) | 228 (63%) | 251 (69%) | 228 (63%) |  |  |
| ***Triage priority*** |  |  |  |  | >0.9 |  |
| Priority 1 | 51 (1%) | 1 (0%) | 1 (0%) | 1 (0%) |  | 0 |
| Priority 2 | 1,269 (17%) | 16 (4%) | 13 (4%) | 16 (4%) |  | 0.1 |
| Priority 3 | 3,225 (42%) | 108 (30%) | 117 (32%) | 108 (30%) |  | -0.04 |
| Priority 4 | 2,897 (38%) | 228 (63%) | 222 (61%) | 228 (63%) |  | 0.01 |
|  | Control, N = 7590 (95.4%)<sup>1</sup> | GEM, N = 362 (4.55%)<sup>1</sup> | Control, N = 362 (50.0%)<sup>1</sup> | GEM, N = 362 (50.0%)<sup>1</sup> |  |  |
| Priority 5 | 148 (2%) | 9 (2%) | 9 (2%) | 9 (2%) |  | 0 |
| *Provenance* |  |  |  |  | >0.9 |  |
| Home | 5,544 (73%) | 125 (35%) | 122 (34%) | 125 (35%) |  | 0.01 |
| Retirement home | 1,651 (22%) | 235 (65%) | 237 (65%) | 235 (65%) |  | 0 |
| Hospital transfer | 110 (1%) | 1 (0%) | 1 (0%) | 1 (0%) |  | 0 |
| Medical clinic | 155 (2%) | 1 (0%) | 2 (1%) | 1 (0%) |  | -0.3 |
| Other | 130 (2%) | 0 (0%) | 0 (0%) | 0 (0%) |  | 0 |
| *Weekday or weekend* |  |  |  |  | >0.9 | 0 |
| Weekday | 5,612 (74%) | 327 (90%) | 327 (90%) | 327 (90%) |  |  |
|  | Control, N = 7590 (95.4%)<sup>1</sup> | GEM, N = 362 (4.55%)<sup>1</sup> | Control, N = 362 (50.0%)<sup>1</sup> | GEM, N = 362 (50.0%)<sup>1</sup> |  |  |
| Weekend | 1,978 (26%) | 35 (10%) | 35 (10%) | 35 (10%) |  |  |
| **Discharge time at index visit** |  |  |  |  | 0.071 | 0.07 |
| After 6PM up to 8AM | 2,927 (39%) | 111 (31%) | 135 (37%) | 111 (31%) |  |  |
| Between 8AM and 6PM | 4,663 (61%) | 251 (69%) | 227 (63%) | 251 (69%) |  |  |
| ***Major diagnostic category*** |  |  |  |  | >0.9 |  |
| Circulatory system diseases | 659 (9%) | 14 (4%) | 18 (5%) | 14 (4%) |  | -0.13 |
| Digestive system diseases | 439 (6%) | 7 (2%) | 10 (3%) | 7 (2%) |  | -0.18 |
| Endocrine, nutritional and metabolic diseases | 101 (1%) | 4 (1%) | 4 (1%) | 4 (1%) |  | 0 |
| Genitourinary system diseases | 197 (3%) | 6 (2%) | 2 (1%) | 6 (2%) |  | 0.5 |
|  | Control, N = 7590 (95.4%)<sup>1</sup> | GEM, N = 362 (4.55%)<sup>1</sup> | Control, N = 362 (50.0%)<sup>1</sup> | GEM, N = 362 (50.0%)<sup>1</sup> |  |  |
| Infectious and parasitic diseases | 171 (2%) | 7 (2%) | 5 (1%) | 7 (2%) |  | 0.16 |
| Mental disorders | 258 (3%) | 49 (14%) | 51 (14%) | 49 (14%) |  | -0.02 |
| Musculoskeletal system and connective tissue diseases | 331 (4%) | 20 (6%) | 23 (6%) | 20 (6%) |  | -0.07 |
| Nervous system diseases | 161 (2%) | 0 (0%) | 0 (0%) | 0 (0%) |  | 0 |
| Respiratory system diseases | 469 (6%) | 25 (7%) | 25 (7%) | 25 (7%) |  | 0 |
| Traumatic injuries | 1,081 (14%) | 145 (40%) | 141 (39%) | 145 (40%) |  | 0.01 |
| Other | 3,723 (49%) | 85 (23%) | 83 (23%) | 85 (23%) |  | 0.01 |
| **Primary complaint or diagnosis** |  |  |  |  | 0.2 |  |
| Cardiovascular and cerebrovascular complaints and | 1,431 (19%) | 20 (6%) | 25 (7%) | 20 (6%) |  | -0.11 |
|  | Control, N = 7590 (95.4%)<sup>1</sup> | GEM, N = 362 (4.55%)<sup>1</sup> | Control, N = 362 (50.0%)<sup>1</sup> | GEM, N = 362 (50.0%)<sup>1</sup> |  |  |
| diagnoses |  |  |  |  |  |  |
| Confusion or delirium | 260 (3%) | 53 (15%) | 46 (13%) | 53 (15%) |  | 0.07 |
| Edema, trouble walking, or joint pain | 333 (4%) | 20 (6%) | 24 (7%) | 20 (6%) |  | -0.09 |
| Falls, syncope, vertigo, or weakness | 511 (7%) | 23 (6%) | 9 (2%) | 23 (6%) |  | 0.43 |
| Fever or hypothermia | 215 (3%) | 5 (1%) | 7 (2%) | 5 (1%) |  | -0.16 |
| Gastrointestinal problems | 621 (8%) | 12 (3%) | 16 (4%) | 12 (3%) |  | -0.14 |
| Respiratory problems | 798 (11%) | 33 (9%) | 37 (10%) | 33 (9%) |  | -0.06 |
| Trauma, fractures, or lesions | 1,090 (14%) | 147 (41%) | 136 (38%) | 147 (41%) |  | 0.04 |
| Urinary tract infections or urinary retention | 104 (1%) | 5 (1%) | 2 (1%) | 5 (1%) |  | 0.43 |
|  | Control, N = 7590 (95.4%)<sup>1</sup> | GEM, N = 362 (4.55%)<sup>1</sup> | Control, N = 362 (50.0%)<sup>1</sup> | GEM, N = 362 (50.0%)<sup>1</sup> |  |  |
| Other | 2,227 (29%) | 44 (12%) | 60 (17%) | 44 (12%) |  | -0.15 |

<sup>1</sup> Mean (SD); n (%)
<sup>2</sup> Two Sample t-test; Fisher's exact test; Fisher's Exact Test for Count Data with simulated p-value (based on 2000 replicates)
<sup>3</sup> Matched variables are italicized
Abbreviations: GEM: Geriatric Emergency Management; SMD: standardized mean difference (continuous variables); RD: risk difference (absolute; categorical values)

We conducted a Cox proportional-hazards regression analysis with the effect of the GEM nurse as the sole predictor variable, having already controlled for covariates with matching. The assumption of proportional hazards was met (χ^2^ = 0.14, *p* = .71). The GEM nurse intervention was associated with a 6% reduction in the risk of returning to the ED within 30 days, but this association was not statistically significant (Hazard Ratio (HR) = 0.94, 95% CI = [0.70, 1.26], *p* = .692). Figure 2 presents survival curves and a risk table with ten-day intervals. The unadjusted estimate without matched cohorts demonstrated that patients seen by the GEM nurse were at a higher risk of ED revisit than all older adult patients visiting the ED (HR = 1.36, 95% CI = [1.1, 1.7], *p* = .005).

**Figure 2.**
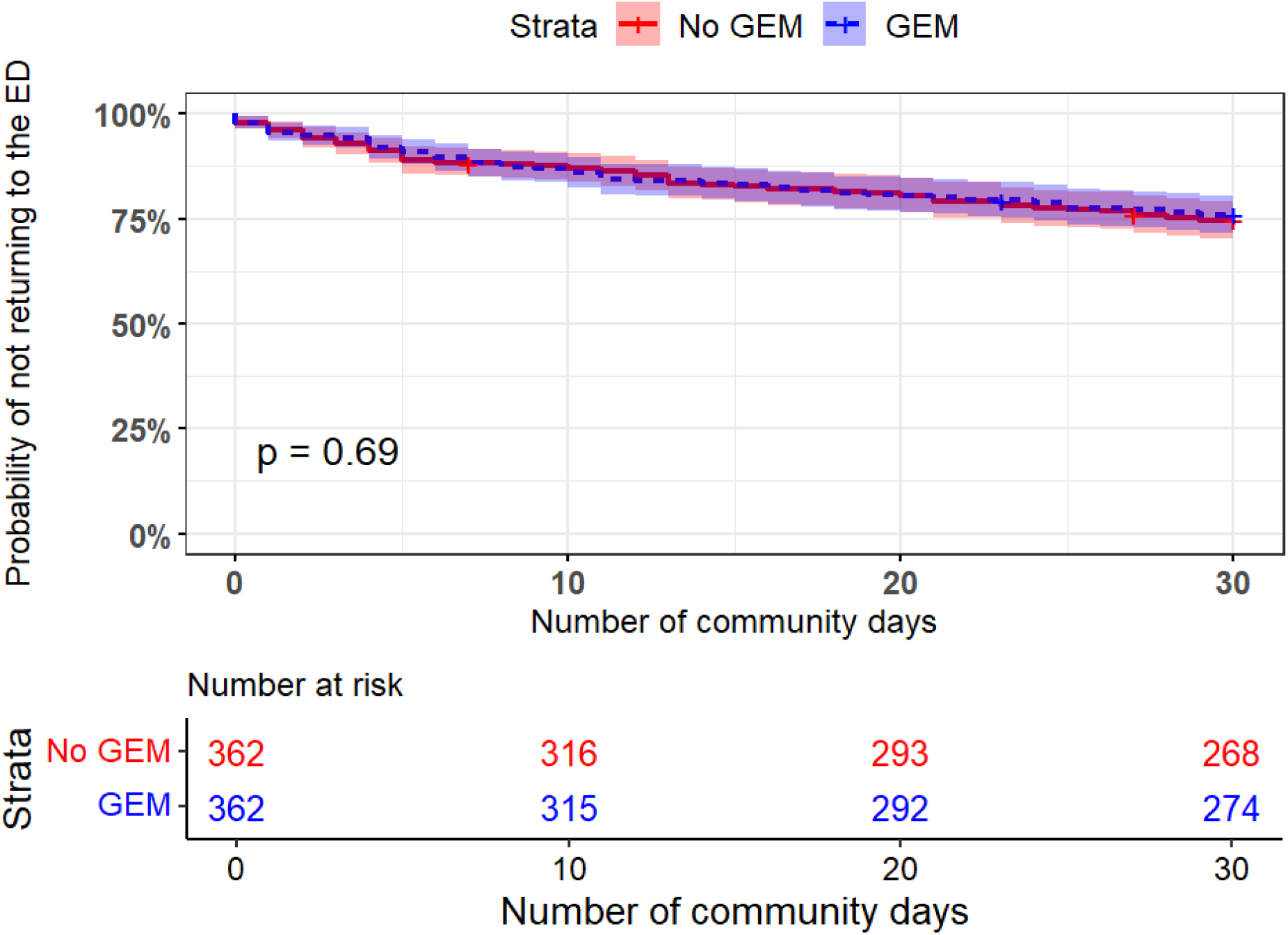
Survival curves and risk table for early revisits to the ED among the propensity-matched sample. The *y*-axis is the probability of success (not returning to the ED), and the *x*-axis is the number of days spent in community after ED discharge. The red solid line denotes the survival curve of the control group, and the blue dotted line the curve of the GEM nurse group. Plus marks (+) indicate censors. Shaded areas denote the 95% confidence intervals.

## Discussion

### Interpretation of Findings

We used a propensity-score matched design to assess the impact of implementing a GEM nurse model in our local ED, controlling for baseline factors related to both being an ideal candidate for the intervention, and the risk of an early ED revisit. We found that—contrary to our hypothesis—the GEM nurse intervention was not associated with a significant decrease in the risk of thirty-day revisits to the ED. The wide confidence intervals around our estimate of a 6% lesser risk of revisit indicate that the risk of revisiting within thirty days associated with the GEM nurse intervention may be reduced by 29.3% but also increased by up to 26.3% relative to the control group.

### Comparison to Previous Studies

Our results align with the conflicting literature on geriatric-focused nurse assessments and interventions in the ED [4,22]. The impact of such assessments and interventions may not impact hospitalizations, re-admissions, or ED revisits at thirty days—and in some cases—an increase in ED revisits after thirty days was observed [23]. Although several studies also reported improvements in revisits and admissions [24]. In a recent umbrella review, the evidence for ED interventions in improving patient experience and quality of life was considered to be low quality, but the effect on reducing ED revisits had considerable variability, ranging from very low to moderate [25]. In a 2024 study on transitional care teams, the thirty-day revisit rate was stable at 14% and was not affected by the transitional care team, and 71% of revisits were due to care needs unrelated to the index visit [26]. Our revisit rate of 24.7% mirrors rates from other sites, but we suspect that thirty-day revisits may not be an adequate outcome measure for assessing the impact of a GEM nurse intervention [27].

### Strengths and Limitations

Strengths include the rigorous use of propensity matching to enhance a quasi-experimental design with adherence to reporting guidelines, as part of a pragmatic approach within a learning health system [28], and is timely given the recognized importance of GEM nurses, evidenced by their permanent implementation in several sites within our catchment area.

The study has several limitations, including its retrospective and monocentric design, reliance on a single GEM nurse, and the inability to perform propensity matching on both geriatric syndrome presentations and major diagnostic criteria. For instance, some patients presented with issues touching upon multiple systems but could only be assigned one major diagnostic category within the database at the ED. The final diagnosis within this database was frequently a primary complaint without a clear diagnosis (e.g., vertigo) and as such, these complaints required extensive cleaning and grouping. These challenges with major diagnostic criteria and primary complaints result from known limitations of administrative data. Most critically, we were unable to account for the added complexity of multiple comorbidities, terminal conditions and frailty, which are characteristics probably immutable by the GEM nurse intervention but are likely drivers of an ED revisit. A measure of frailty—a potential source of residual confounding in our cohort—will be important to include to measure the impact of future GEM nurse interventions [29].

### Health System Implications

The most effective discharge interventions extend beyond referral and use a clinical risk prediction tool to identify those who would most benefit from the intervention [30]. Clinical risk assessments using tools such as InterRAI ED Screener [31] or the Identification of Seniors at Risk (ISAR) [32] were not performed systematically in this study, but the GEM nurse did use a personalized structured assessment form that included screening for delirium using the Confusion Assessment Method, [33] functional decline, immobilization, falls, wound screening, dehydration, and incontinence (see Appendix A). Telephone follow-up has previously been efficacious [34]. Follow-up has been identified by both patients and their caregivers as an element needing immediate improvement to ameliorate the quality of care transitions from the ED [35,36]. For nursing interventions, there is conflicting moderate but inconsistent agreement across the studies for the effectiveness of nurse-led interventions [23]. Generally, the main weakness of these interventions arise from the fact that they do not extend past the ED to the community. Even so, GEM nurses are considered the cornerstone of geriatric emergency care and are thought to improve the quality of care, accessibility, and quality of life within our province [37]. In 2021, the Quebec ministry of health and social services recommended the implementation of a GEM nurse in every ED in the province [38].

### Research Implications

A challenge to contend with in future work is the over-reliance on using service-level measures (e.g., revisits, admissions, length of stay) to assess the efficacy of interventions at the ED [39]. Current protocols and investigations appear to be shifting towards evaluating quality of life and symptom burden as primary outcome variables while keeping service-level measures as secondary outcomes [40]. In recent and ongoing trials, both service-level measures and qualitative interviews of patients, caregivers, and treating physicians were triangulated in the assessment of the intervention [26].

## Conclusion

The GEM nurse intervention did not significantly reduce thirty-day revisits to the ED among older adult patients matched on baseline characteristics, although we were unable to match for other relevant psycho-social and clinical characteristics, such as frailty, terminal conditions, or caregiver burden. While GEM nurses are an established and important role in ED settings, our results suggest that the precise components of a GEM nurse intervention merit further investigation due to the highly complex and personalized nature of the intervention.

## Acknowledgements

We acknowledge the invaluable support of participating patients and their caregivers. We also thank Denis Roy, Lise Lavoie, Maryse Turcotte, Josée Chouinard, Don Melady, Samir Sinha, Marie-Soleil Hardy, Richard Fleet, Annie LeBlanc, Marcel Émond, Jean-Louis Denis, Éric Mercier, Valérie Roy, Rosalie Beaudoin, Julie Émond, Marie-Josée Sirois, Isabelle Pelletier, Marie-Hélène Savard, Vanessa Couture, Raphaëlle Giguère, Sam Chandavong, Lyna Abrougui, El Kebir Ghandour, Daniel Paré, Tom van de Belt and David Buckeridge for their support and expertise in planning and contributing to the LEARNING WISDOM project. Data and analysis scripts are not publicly available but may be requested by contacting the corresponding author.

## Ethics approval

The protocol for this study was approved by the Centre intégré de santé et de services sociaux - Chaudière Appalaches (CISSS-CA, Québec, Canada) Ethics Review Committee (project #2018-462, 2018-007).

## Data availability

The anonymized data, analytic methods and materials are available to other researchers for replication purposes, upon reasonable request to the corresponding author. The analysis methods for this specific study were not preregistered but are nested within the protocol of a mixed methods implementation study (LEARNING WISDOM, ClinicalTrials.gov ID: NCT04093245). That protocol can be found here: https://www.researchprotocols.org/2020/8/e17363.

## References

1. Stolee P, Elliott J, Giguere AM, Mallinson S, Rockwood K, Sims Gould J, et al. Transforming primary care for older Canadians living with frailty: mixed methods study protocol for a complex primary care intervention. BMJ Open. 2021;11:e042911.

2. Gruneir A, Silver MJ, Rochon PA. Emergency department use by older adults: a literature review on trends, appropriateness, and consequences of unmet health care needs. Med Care Res Rev. 2011;68:131–55.

3. McCusker J, Verdon J. Do geriatric interventions reduce emergency department visits? A systematic review. J Gerontol A Biol Sci Med Sci. 2006;61:53–62.

4. Leaker H, Fox L, Holroyd-Leduc J. The Impact of Geriatric Emergency Management Nurses on the Care of Frail Older Patients in the Emergency Department: a Systematic Review. Can Geriatr J [Internet]. 2020 [cited 2024 Jul 10];23:250–6. Available from: https://www.ncbi.nlm.nih.gov/pmc/articles/PMC7458600/

5. Miller DK, Lewis LM, Nork MJ, Morley JE. Controlled trial of a geriatric case-finding and liaison service in an emergency department. J Am Geriatr Soc. 1996;44:513–20.

6. Mion LC, Palmer RM, Meldon SW, Bass DM, Singer ME, Payne SMC, et al. Case finding and referral model for emergency department elders: a randomized clinical trial. Ann Emerg Med. 2003;41:57–68.

7. McCusker J, Dendukuri N, Tousignant P, Verdon J, Poulin de Courval L, Belzile E. Rapid Two-stage Emergency Department Intervention for Seniors: Impact on Continuity of Care. Academic Emergency Medicine [Internet]. 2003 [cited 2024 Oct 8];10:233–43. Available from: https://onlinelibrary.wiley.com/doi/abs/10.1197/aemj.10.3.233

8. Davis RA, Dinh MM, Bein KJ, Veillard A-S, Green TC. Senior work-up assessment and treatment team in an emergency department: a randomised control trial. Emerg Med Australas. 2014;26:343–9.

9. Wright PN, Tan G, Iliffe S, Lee D. The impact of a new emergency admission avoidance system for older people on length of stay and same-day discharges. Age and Ageing [Internet]. 2014 [cited 2024 Jul 10];43:116–21. Available from: 10.1093/ageing/aft086

10. Shanley C, Sutherland S, Tumeth R, Stott K, Whitmore E. Caring for the Older Person in the Emergency Department: The ASET Program and the Role of the ASET Clinical Nurse Consultant in South Western Sydney, Australia. Journal of Emergency Nursing [Internet]. 2009 [cited 2024 Jul 10];35:129–33. Available from: https://www.jenonline.org/article/S0099-1767(08)00290-0/abstract

11. Hwang U, Dresden SM, Rosenberg MS, Garrido MM, Loo G, Sze J, et al. Geriatric Emergency Department Innovations: Transitional Care Nurses and Hospital Use. Journal of the American Geriatrics Society. 2018;66:459–66.

12. Yao XI, Wang X, Speicher PJ, Hwang ES, Cheng P, Harpole DH, et al. Reporting and Guidelines in Propensity Score Analysis: A Systematic Review of Cancer and Cancer Surgical Studies. J Natl Cancer Inst. 2017;109:djw323.

13. von Elm E, Altman DG, Egger M, Pocock SJ, Gøtzsche PC, Vandenbroucke JP, et al. The Strengthening the Reporting of Observational Studies in Epidemiology (STROBE) statement: guidelines for reporting observational studies. J Clin Epidemiol. 2008;61:344–9.

14. Archambault PM, Rivard J, Smith PY, Sinha S, Morin M, LeBlanc A, et al. Learning Integrated Health System to Mobilize Context-Adapted Knowledge With a Wiki Platform to Improve the Transitions of Frail Seniors From Hospitals and Emergency Departments to the Community (LEARNING WISDOM): Protocol for a Mixed-Methods Implementation Study. JMIR Res Protoc. 2020;9:e17363.

15. Ghandour EK, Leblond S, Binette S, Rivard J, Joanisse J, Carreau L, et al. Implementation of the Acute Care for Elders Strategy to Improve the Quality of Care Transitions in Quebec and Ontario: a Retrospective Multiple Case Study. Canadian geriatrics journal : CGJ [Internet]. 2023 [cited 2024 Oct 8];26. Available from: https://pubmed.ncbi.nlm.nih.gov/38045881/

16. Shukla DM, Faber EB, Sick B. Defining and Characterizing Frequent Attenders: Systematic Literature Review and Recommendations. J Patient Cent Res Rev [Internet]. 2020 [cited 2024 Oct 8];7:255–64. Available from: https://www.ncbi.nlm.nih.gov/pmc/articles/PMC7398628/

17. Ho D, Imai K, King G, Stuart EA. MatchIt: Nonparametric Preprocessing for Parametric Causal Inference. Journal of Statistical Software [Internet]. 2011 [cited 2024 Sep 11];42:1–28. Available from: 10.18637/jss.v042.i08

18. Gu XS, Rosenbaum PR. Comparison of Multivariate Matching Methods: Structures, Distances, and Algorithms. Journal of Computational and Graphical Statistics [Internet]. 1993 [cited 2024 Sep 13];2:405–20. Available from: https://www.tandfonline.com/doi/abs/10.1080/10618600.1993.10474623

19. Simoneau G, Mitroiu M, Debray TP, Wei W, Wijn SR, Magalhães JC, et al. Visualizing the target estimand in comparative effectiveness studies with multiple treatments. J Comp Eff Res [Internet]. 2024 [cited 2024 Sep 11];13:e230089. Available from: https://www.ncbi.nlm.nih.gov/pmc/articles/PMC10842272/

20. Austin PC. An Introduction to Propensity Score Methods for Reducing the Effects of Confounding in Observational Studies. Multivariate Behav Res [Internet]. 2011 [cited 2024 Sep 11];46:399–424. Available from: https://www.ncbi.nlm.nih.gov/pmc/articles/PMC3144483/

21. Beveridge R, Ducharme J, Janes L, Beaulieu S, Walter S. Reliability of the Canadian emergency department triage and acuity scale: interrater agreement. Ann Emerg Med. 1999;34:155–9.

22. Memedovich A, Asante B, Khan M, Eze N, Holroyd BR, Lang E, et al. Strategies for improving ED-related outcomes of older adults who seek care in emergency departments: a systematic review. Int J Emerg Med. 2024;17:16.

23. Malik M, Moore Z, Patton D, O’Connor T, Nugent LE. The impact of geriatric focused nurse assessment and intervention in the emergency department: A systematic review. Int Emerg Nurs. 2018;37:52–60.

24. Aldeen AZ, Courtney DM, Lindquist LA, Dresden SM, Gravenor SJ. Geriatric emergency department innovations: preliminary data for the geriatric nurse liaison model. J Am Geriatr Soc. 2014;62:1781–5.

25. Conneely M, Leahy S, Dore L, Trépel D, Robinson K, Jordan F, et al. The effectiveness of interventions to reduce adverse outcomes among older adults following Emergency Department discharge: umbrella review. BMC Geriatrics [Internet]. 2022 [cited 2024 Oct 3];22:462. Available from: 10.1186/s12877-022-03007-5

26. Pepping RMC, Vos RC, Numans ME, Kroon I, Rappard K, Labots G, et al. An emergency department transitional care team prevents unnecessary hospitalization of older adults: a mixed methods study. BMC Geriatr. 2024;24:668.

27. Schouten B, Driesen BEJM, Merten H, Burger BHCM, Hartjes MG, Nanayakkara PWB, et al. Experiences and perspectives of older patients with a return visit to the emergency department within 30 days: patient journey mapping. Eur Geriatr Med [Internet]. 2022 [cited 2024 Jun 14];13:339–50. Available from: https://www.ncbi.nlm.nih.gov/pmc/articles/PMC9018642/

28. Menear M, Blanchette M-A, Demers-Payette O, Roy D. A framework for value-creating learning health systems. Health Research Policy and Systems [Internet]. 2019 [cited 2024 Oct 17];17:79. Available from: 10.1186/s12961-019-0477-3

29. Muscedere J, Bagshaw SM, Kho M, Mehta S, Cook DJ, Boyd JG, et al. Frailty, Outcomes, Recovery and Care Steps of Critically Ill Patients (FORECAST): a prospective, multi-centre, cohort study. Intensive Care Med [Internet]. 2024 [cited 2024 Oct 4];50:1064–74. Available from: 10.1007/s00134-024-07404-9

30. Karam G, Radden Z, Berall LE, Cheng C, Gruneir A. Efficacy of emergency department-based interventions designed to reduce repeat visits and other adverse outcomes for older patients after discharge: A systematic review. Geriatr Gerontol Int. 2015;15:1107–17.

31. Mowbray FI, Heckman G, Hirdes JP, Costa AP, Beauchet O, Eagles D, et al. Examining the utility and accuracy of the interRAI Emergency Department Screener in identifying high-risk older emergency department patients: A Canadian multiprovince prospective cohort study. Journal of the American College of Emergency Physicians Open [Internet]. 2023 [cited 2024 Oct 17];4:e12876. Available from: https://onlinelibrary.wiley.com/doi/abs/10.1002/emp2.12876

32. McCusker J, Warburton RN, Lambert SD, Belzile E, de Raad M. The Revised Identification of Seniors At Risk screening tool predicts readmission in older hospitalized patients: a cohort study. BMC Geriatrics [Internet]. 2022 [cited 2024 Oct 17];22:888. Available from: 10.1186/s12877-022-03458-w

33. Wei LA, Fearing MA, Sternberg EJ, Inouye SK. The Confusion Assessment Method (CAM): A Systematic Review of Current Usage. Journal of the American Geriatrics Society [Internet]. 2008 [cited 2024 Oct 17];56:823. Available from: https://pmc.ncbi.nlm.nih.gov/articles/PMC2585541/

34. Lowthian JA, McGinnes RA, Brand CA, Barker AL, Cameron PA. Discharging older patients from the emergency department effectively: a systematic review and meta-analysis. Age and Ageing [Internet]. 2015 [cited 2024 Jul 10];44:761–70. Available from: 10.1093/ageing/afv102

35. Couture V, Germain N, Côté É, Lavoie L, Robitaille J, Morin M, et al. Transitions of care for older adults discharged home from the emergency department: an inductive thematic content analysis of patient comments. BMC Geriatr [Internet]. 2024 [cited 2024 Jan 29];24:8. Available from: 10.1186/s12877-023-04482-0

36. Germain N, Jémus-Gonzalez E, Couture V, Côté É, Morin M, Toulouse-Fournier A, et al. Caregivers’ burden of care during emergency department care transitions among older adults: a mixed methods cohort study. BMC Geriatrics [Internet]. 2024 [cited 2024 Oct 9];24:788. Available from: 10.1186/s12877-024-05388-1

37. Poulin V, Mailhot-Bisson D, Turcotte-Brousseau A-A. Le déploiement du rôle d’une infirmière en pratique avancée en gériatrie à l’urgence : une innovation en Estrie. soinsurgence [Internet]. 2021 [cited 2024 Oct 3];2:35–44. Available from: https://www.erudit.org/fr/revues/soinsurgence/2021-v2-n2-soinsurgence08256/1101813ar/

38. Brousseau A-A, Grégoire M. Vers un service d’urgence adapté pour la personne âgée - Cadre de référence [Internet]. Ministère de la Santé et des Services sociaux, editor. Québec; 2021 [cited 2024 Oct 3]. Available from: https://numerique.banq.qc.ca/patrimoine/details/52327/4501881

39. Preston L, Oppen JD van, Conroy SP, Ablard S, Woods HB, Mason SM. Improving outcomes for older people in the emergency department: a review of reviews. Emerg Med J [Internet]. 2021 [cited 2024 Oct 3];38:882–8. Available from: https://emj.bmj.com/content/38/12/882

40. Koirala B, Badawi S, Frost S, Ferguson C, Hager DN, Street L, et al. Study protocol for Care cOORDInatioN And sympTom managEment (COORDINATE) programme: a feasibility study. BMJ Open [Internet]. 2023 [cited 2024 Oct 3];13:e072846. Available from: https://bmjopen.bmj.com/content/13/12/e072846

