## Supplement for "Impact of a geriatric emergency management nurse on thirty-day emergency department revisits: a propensity score matched case-control study"

### **Appendix A. Evaluation form used by the Geriatric Emergency Nurse (GEM-N): English translation**

Patients were not randomized to the GEM nurse intervention. Instead, the intervention was focused on a pragmatic approach of targeting a specific phenotype in older adult patients that would benefit most from a GEM nurse consultation and evaluation and would improve ED flow. Patients were identified by the GEM nurse each morning by attending the daily team brief led by the ED charge nurse, and by screening the list of patients cared for in the ED observation unit. ED physicians could also refer patients to the GEM nurse. The GEM nurse was available during standard weekday working hours, excluding holidays.

The targeted geriatric assessment included a revision of current and past patient files, and a collection of information from the patient and validation of that information with ED staff, staff at the patient’s care home or community services center, and with family. This consultation also consisted of: 1) a physical exam with particular attention paid to mobility, autonomy at home, activities of daily living, and mental state; 2) identification of risk factors for certain geriatric syndromes and patient needs; 3) the construction of an intervention plan designed to prevent falls and/or alleviate geriatric syndromes; and 4) to develop a care transition plan by organizing home care services or convalescent care, and to educate both the patient and their caregivers on the results of the consultation and on the community resources at their disposal.

**Please contact the corresponding author for the original French version.**

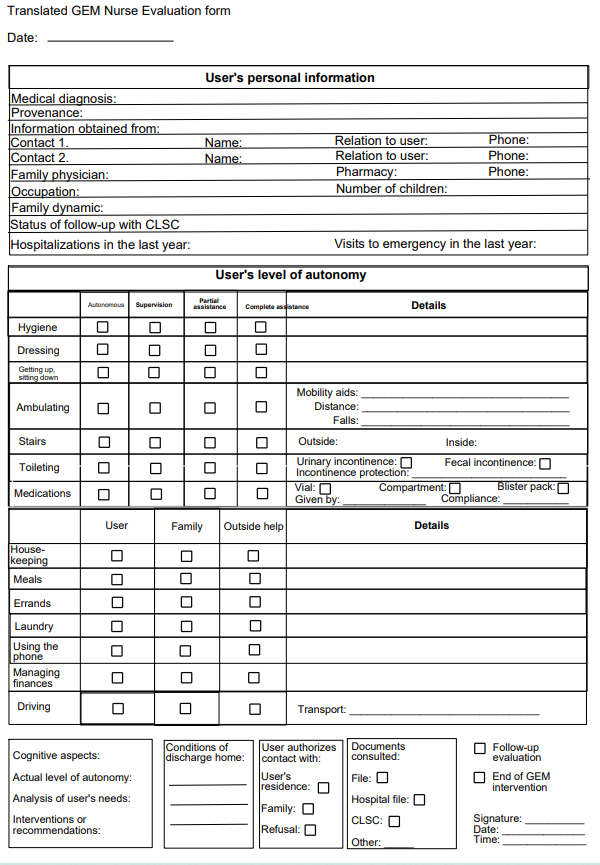

### **Appendix B. Propensity score checklist from Yao et al., 2017**

| Section/topic | Item | No.[^*^](https://www.ncbi.nlm.nih.gov/pmc/articles/PMC6059208/table/djw323-T4/?report=objectonly#tblfn11) | Recommendation |
| --- | --- | --- | --- |
| **Title and abstract** | ✓ | 1 | Indicate the use of propensity analysis with a commonly used term in the title or the abstract |
| **Methods** |  |  |  |
| Bias | ✓ | 9 | Describe how propensity score analysis was used to address bias |
| Statistical analyses | ✓ | 12 | Describe all the analytic methods, including the propensity score methods, eg, PSM, PSW, PSS, CAPS |
|  | ✓ | 13 | Indicate the model used to estimate propensity score |
|  | ✓ | 14 | State the variables included in the propensity score model |
|  | ✓ | 15 | Explain the variable selection procedure for propensity score model |
|  | ✓ | 16 | PSM: Explicitly state the matching algorithm and distance metric, indicate matching ratio (1:m matching), indicate whether sampling with or without replacement was used, describe the statistical methods for the analysis of matched data, report the package used to create matched sample, and describe methods for assessing the comparability of baseline characteristics in the matched groups |
|  | ✓ | 17 | PSW: Describe methods for assessing the comparability of baseline characteristics in the weighted groups |
|  | ✓ | 18 | PSS: Give the number of strata and describe methods for assessing the comparability of baseline characteristics in each stratum |
|  | ✓ | 19 | Explain how assumption of propensity score analysis was examined |
|  | ✓ | 20 | Explain how missing data in propensity score estimation were addressed |
| **Results** |  |  |  |
| Participants | ✓ | 25.4 | PSM: Report the sample size for each treatment group before and after matching |
| Patient characteristics | ✓ | 28 | Describe the distribution of baseline characteristics for each group before propensity score analysis |
|  | ✓ | 29 | PSM, PSW, PSS: Describe the distribution of baseline characteristics in the matched/weighted groups or in each stratum, and describe the results of the comparability of baseline characteristics |
|  | ✓ | 30 | Indicate number of patients with missing data for each variable of interest, especially the variables used in propensity score model |
| Main results | ✓ | 32 | Give propensity score analysis estimates and their precision, eg, 95% confidence interval |
|  | ✓ | 33 | If applicable, give unadjusted estimates and/or adjusted estimates and their precision, eg, 95% confidence interval, and make clear which additional factors were adjusted for |
| **Discussion** |  |  | |
| Interpretation | ✓ | 38 | Discuss whether imbalance of baseline characteristics still exists, and give a cautious interpretation |
| Generalizability | ✓ | 40 | PSM: Discuss the possibility and potential influence of incomplete matching, especially the studies in which the matched sample size is less than 50% |

### **Appendix C. STROBE checklist for case control studies from von Elm et al. (2007)**

|  | **Item No** | **Recommendation** |
| --- | --- | --- |
| **Title and abstract** | 1 | (*a*) Indicate the study’s design with a commonly used term in the title or the abstract ✓ |
|  |  | (*b*) Provide in the abstract an informative and balanced summary of what was done and what was found ✓ |
| **Introduction** | | |
| Background/rationale | 2 | Explain the scientific background and rationale for the investigation being reported ✓ |
| Objectives | 3 | State specific objectives, including any prespecified hypotheses ✓ |
| **Methods** | | |
| Study design | 4 | Present key elements of study design early in the paper ✓ |
| Setting | 5 | Describe the setting, locations, and relevant dates, including periods of recruitment, exposure, follow-up, and data collection ✓ |
| Participants | 6 | (*a*) Give the eligibility criteria, and the sources and methods of case ascertainment and control selection. Give the rationale for the choice of cases and controls ✓ |
|  |  | (*b*) For matched studies, give matching criteria and the number of controls per case ✓ |
| Variables | 7 | Clearly define all outcomes, exposures, predictors, potential confounders, and effect modifiers. Give diagnostic criteria, if applicable ✓ |
| Data sources/ measurement | 8* | For each variable of interest, give sources of data and details of methods of assessment (measurement). Describe comparability of assessment methods if there is more than one group ✓ |
| Bias | 9 | Describe any efforts to address potential sources of bias ✓ |
| Study size | 10 | Explain how the study size was arrived at ✓ |
| Quantitative variables | 11 | Explain how quantitative variables were handled in the analyses. If applicable, describe which groupings were chosen and why ✓ |
| Statistical methods | 12 | (*a*) Describe all statistical methods, including those used to control for confounding ✓ |
|  |  | (*b*) Describe any methods used to examine subgroups and interactions ✓ |
|  |  | (*c*) Explain how missing data were addressed ✓ |
|  |  | (*d*) If applicable, explain how matching of cases and controls was addressed ✓ |
|  |  | (*e*) Describe any sensitivity analyses ✓ |
| **Results** | | |
| Participants | 13* | (a) Report numbers of individuals at each stage of study—eg numbers potentially eligible, examined for eligibility, confirmed eligible, included in the study, completing follow-up, and analysed ✓ |
|  |  | (b) Give reasons for non-participation at each stage ✓ |
|  |  | (c) Consider use of a flow diagram ✓ |
| Descriptive data | 14* | (a) Give characteristics of study participants (eg demographic, clinical, social) and information on exposures and potential confounders ✓ |
|  |  | (b) Indicate number of participants with missing data for each variable of interest ✓ |
| Outcome data | 15* | Report numbers in each exposure category, or summary measures of exposure ✓ |
| Main results | 16 | (*a*) Give unadjusted estimates and, if applicable, confounder-adjusted estimates and their precision (eg, 95% confidence interval). Make clear which confounders were adjusted for and why they were included ✓ |
|  |  | (*b*) Report category boundaries when continuous variables were categorized ✓ |
|  |  | (*c*) If relevant, consider translating estimates of relative risk into absolute risk for a meaningful time period ✓ |
